# Maternal diabetes influences neonatal obesity-adiposity but not in later life Offspring obesity in diabetic pregnancy

**DOI:** 10.1101/2023.04.28.23289246

**Authors:** Sayali S. Deshpande-Joshi, Sonali S. Wagle, Madhura K. Deshmukh, Hemant S. Damle, Suhas R. Otiv, Sanat B. Phatak, Smita N. Dhadge, Shubha S. Ambardekar, Dattatray S. Bhat, Deepa A. Raut, Rajashree P. Kamat, Sayali G. Wadke, Kalyanaraman Kumaran, Giriraj R. Chandak, Chittaranjan S. Yajnik

## Abstract

**Background:** Based on studies in overweight-obese populations, it is tacitly assumed that maternal hyperglycemia is responsible for obesity-adiposity at birth and in later life.

**Study design:** Two hospital based case control studies: 1) Neonatal outcomes, 2) Later life outcomes.

**Methods:** We studied associations of neonatal and later life obesity-adiposity [age and sex-adjusted BMI, waist circumference, skinfolds, and body fat percent by Dual energy X-ray Absorptiometry (DXA)] in offspring of mothers with diabetes (ODM) and those of mothers without diabetes (ONDM). Exposures were parental hyperglycemia and overweight-obesity.

**Results:** Neonatal study included 372 non-diabetic and 816 diabetic pregnancies [74 type 1 diabetes, 102 type 2 diabetes, 640 gestational diabetes (GDM)]. Mothers with type 1 diabetes were the youngest, thinnest, and with highest HbA1c. Maternal glycemia but not BMI was associated with neonatal obesity-adiposity. Thus, neonates of mothers with type 1 diabetes had highest ponderal index, abdominal circumference, and skinfolds.

Later life study included 200 ODM (25 type 1 diabetes, 22 type 2 diabetes, 153 GDM) and 177 age, sex and socio-economic matched ONDM (2 to 26 y). Their obesity-adiposity was associated with bi-parental overweight-obesity in an additive manner, but not with parental diabetes. Offspring birth weight was also positively associated. Offspring of mothers with type 1 diabetes had the lowest and offspring of mothers with type 2 diabetes the highest obesity-adiposity.

**Conclusion:** Neonatal obesity-adiposity is driven by maternal glycemia while later life obesity-adiposity by bi-parental obesity. Our results provide a clear insight into pathogenesis of obesity-adiposity in the offspring.

**Article Highlights:** It is tacitly assumed that maternal diabetes is responsible for offspring obesity-adiposity.

We examined the determinants of obesity-adiposity in intrauterine and in later life in children born to mothers with type 1, type 2 and GDM. Paternal influence was also investigated.

Mothers with type 1 diabetes were the thinnest and most hyperglycemic. Their children were the most obese-adipose at birth but thinnest in later life. Later life obesity-adiposity was driven by bi-parental overweight-obesity, not by diabetes.

Our findings suggest that strict maternal metabolic control during pregnancy will reduce macrosomia while targeting obesogenic family environment may reduce later life offspring obesity-adiposity.

---

In 2021, an estimated 16.7% of live births worldwide (> 20 million) were exposed to some form of glucose intolerance in pregnancy. Of these, 10%-20% had pre-gestational diabetes (Type 1 and Type 2, other) while the majority (80%) had gestational diabetes (1). Maternal diabetes during pregnancy is associated with adverse short- and long-term outcomes for the offspring (2). The main short-term effect is excessive fetal growth and its sequelae in the peripartum period (difficult labour, need for interventional delivery, post-natal complications etc.). Pedersen postulated that transfer of excess maternal glucose to the fetus in a diabetic pregnancy stimulates fetal islets, resulting in hyperinsulinemia and macrosomia (3). Freinkel proposed that an excess transfer of a “mixture” of maternal nutrients (glucose, lipids, and amino acids) in diabetic pregnancies promotes fetal macrosomia and long-term risk of obesity and diabetes (fuel mediated teratogenesis) (4). Thus, maternal diabetes has been proposed to contribute to the cascading epidemic of obesity-adiposity and diabetes (5-7).

In the populations from high-income countries, maternal diabetes and obesity are strongly associated, making it difficult to distinguish individual contributions of these two related yet distinct exposures on offspring obesity-adiposity. Current thinking tacitly assumes that maternal ‘diabetes’ influences both intra uterine and later life offspring obesity-adiposity, though some studies have pointed out that maternal obesity may be the actual driver (8, 9). In the classic Pima Indian study, offspring born in diabetic pregnancies were overweight-obese at 5-19 yrs of age compared to those born in non-diabetic pregnancies (10). However, parental obesity was also associated with offspring obesity-adiposity from early childhood to adolescent age, the association became stronger with increasing age and paternal association was stronger than the maternal (11). In another Pima Indian study, siblings exposed to maternal diabetes *in utero* had higher BMI compared to siblings not exposed but no such difference was observed in the offspring born before and after diagnosis of diabetes in fathers (12). This suggested that intrauterine exposure to hyperglycemia is associated with obesity-adiposity apart from genetic effects. However, in this population there is little information on independent effects of parental overweight-obesity and glycemia. In a German study, BMI of both mother and father at the time of follow-up were predictors for an increased BMI of the child in addition to maternal pregnancy glycemia (13). Recent follow-up reports in the Hyperglycemia and Adverse Pregnancy Outcome (HAPO) study are suggestive of an independent effect of maternal diabetes and obesity on offspring obesity-adiposity, higher maternal BMI attenuated the effect of maternal diabetes (14). Interestingly, there was no protection against childhood overweight-obesity in children born to intensively treated GDM mothers in Randomized Controlled Trials (15, 16). Clarity about independent effects of maternal diabetes and obesity may come from studies in populations where these two characteristics are dissociated. Studying paternal contribution would further improve our understanding of these complex inter-generational associations.

GDM is common in India, affecting 10-15% of pregnancies (17), Indian women with GDM are younger and have a lower BMI than European women with GDM (18). At the Diabetes Unit, King Edward Memorial Hospital Research Centre (KEMHRC), Pune we have collected data on substantial numbers of pregnancies with type 1 diabetes, type 2 diabetes and GDM over last three decades. We have observed that mothers with type 1 diabetes are the thinnest and most hyperglycemic, while those with type 2 diabetes and GDM are heavier and less hyperglycemic. This provide an opportunity to investigate disparate associations of maternal glycemia and body size with offspring obesity-adiposity at birth and in later life. We also investigated contribution of fathers to these associations in the later life study.

## Methods

We used data from clinical records and research studies of Diabetes Unit, KEMHRC, Pune, India to investigate determinants of neonatal and later life obesity-adiposity in the offspring born to mothers with and without diabetes.

### Studies and Participants

#### Neonatal body size

The **D**iabetes **I**n **P**regnancy (DIP) study included 816 diabetic pregnancies (type 1 diabetes, type 2 diabetes and GDM) managed from 1986 to 2020. Control data on 372 normal glucose tolerant (NGT) pregnancies came from two observational studies which investigated the association of maternal nutritional and metabolic characteristics with neonatal size: 1) International Atomic Energy Association (IAEA)-B12 study (2003-06) (19) and 2) **In**tergenerational programming of **Dia**besity in offspring of mothers with **G**estational **D**iabetes **M**ellitus, 2014-17) (InDiaGDM).

#### Later life body size and composition

The InDiaGDM study also investigated obesity-adiposity in 200 offspring born to 176 diabetic mothers (24 siblings born in a subsequent diabetic pregnancy) (2-24 y of age) and 177 offspring born in non-diabetic pregnancies (schoolmates or neighbours of ODMs), matched for age, sex and socio-economic status (SES). We studied both mothers and fathers in this study.

### Exposures

In the neonatal study, exposures were maternal diabetes during pregnancy (none, type 1 diabetes, type 2 diabetes and GDM) and maternal BMI at delivery. We considered maternal diabetes type as a surrogate of degree of maternal hyperglycemia: type 1 diabetes being the most hyperglycemic, type 2 diabetes intermediate and GDM the least hyperglycemic. Paternal data was not available.

We obtained information on the type of maternal diabetes, demographics, obstetric history, pregnancy complications, treatment, and delivery details (sex of the neonate, gestational age at delivery, mode of delivery) from the clinical records. Maternal type 1 diabetes and type 2 diabetes were defined by ADA clinical criteria. GDM was diagnosed by a fasting 75 g OGTT (WHO 1999 criteria [FPG ≥ 6.1 mmol/L and/ 2-hr plasma glucose ≥ 7.7 mmol/L] till 2012, and IADPSG criteria thereafter (FPG ≥ 5.1 mmol/L or 2-hr glucose ≥ 8.5 mmol/L).

Mothers with type 1 diabetes were treated with multiple daily doses of insulin, none had used insulin pump. Mothers with type 2 diabetes and GDM were treated with lifestyle advice, and oral anti-diabetic drugs (OADs, metformin or acarbose) and insulin if necessary. Maternal weight, height, waist circumference, bicep, tricep, subscapular and suprailiac skinfold thicknesses were measured by trained staff within 72 hours of delivery using a standard protocol (20) (Supplemental Table S1). Similar information was also available in mothers without diabetes.

In the later life study, exposures included type of maternal diabetes in pregnancy, paternal glucose intolerance and parental overweight-obesity measured at follow-up. Paternal glycemic status was defined as hyperglycemia (yes/no). Hyperglycemic included: known diabetic + prediabetes + diabetes on a 75 g oral glucose tolerance test (OGTT, ADA 2014). Overweight-obesity refers to BMI >=25 kg/m^2^. SES of the family was evaluated using the Standard of Living Index (SLI) tool (National Family Health Survey-2, 1998-99), higher score denotes higher SES. Pubertal staging (Tanner) was done for offspring between 8 to 18 years of age, including date of menarche. Offspring birth weight was obtained either from medical records or by maternal recall.

### Outcomes

We use term overweight-obesity for body size (BMI, ponderal index, abdominal and waist circumference) and adiposity for body fat measurements (skinfolds or DXA).

#### Neonatal

Trained research staff performed detailed anthropometry (birth weight, crown heel length, head circumference, abdominal circumference, triceps and subscapular skinfold thicknesses) within 72 hours of birth using standardized protocols (20) (Supplemental Table S1). Obesity-adiposity were calculated as gestational age and sex specific SD score >85^th^ percentile by residual method for combined ODM and ONDM data. These included obesity (ponderal index), abdominal obesity (abdominal circumference), and adiposity (sum of skinfolds). Small for gestational age (SGA) and large for gestational age (LGA) were defined using INTERGROWTH-21 standards (2014).

#### Later life

We measured weight, height, waist and hip circumferences, bicep, tricep, subscapular and suprailiac skinfold thicknesses by standardized protocol (21) (Supplemental Table S1) and fat and lean mass and bone mineral density by DXA. Inter- and intra-observer variation studies were conducted periodically and training was reinforced to maintain the quality of measurements. The coefficient of variation (CV) between observers for different measurements was less than 3%. Offspring overweight-obesity (BMI) was calculated using IOTF method for 2-18y (22) and WHO criteria for >=18y (23), for both it was >=25 Kg/m^2^. In absence of reference values for central obesity (waist circumference) and adiposity (sum of skinfolds) in this age group, we defined these as age, sex and pubertal stage specific SD score >85^th^ percentile by residual method for combined ODM and ONDM data.

### Laboratory methods

Venous plasma glucose was measured using glucose oxidase-peroxidase method (Hitachi 902, Roche Diagnostics GmbH, Germany). Measurements were subject to external quality control (EQAS) and had CV <5%. HbA1c was measured chromatographically on Biorad D10 machine (Hercules, California).

### Statistical methods

For the neonatal study, power was calculated retrospectively for the outcome of obesity-adiposity. Studying 800 diabetic and 370 NGT pregnancies would provide a power of 90% at 5% significance level for a difference of 0.2 SD for ponderal index between the two groups.

For the later life study, we considered BMI as the primary outcome, using previously reported difference between Indian ODMs and ONDMs (24, 25). In the age group <10 years, 108 participants per group would allow detection of 0.5 kg/m^2^ difference at 90% power and 5% significance level; in those ≥10 years old, 77 participants per group would allow detection of 1.0 kg/m^2^ difference.

Data are presented as mean (SD) for normally distributed variables and as median (25^th^–75^th^ percentile) for skewed variables; skewed data were normalized before analysis by using transformation (log, reciprocal, cube root, square root). Comparisons between means was made using student’s t-test and medians by Mann Whitney’s U-test. ANCOVA was used when adjustments for confounders were necessary. Difference in proportions was tested using the Z-test.

Association between maternal type of diabetes and neonatal obesity-adiposity was examined using multiple logistic regression analysis, using NGT mothers as the reference. Maternal BMI was also included in this model, using lowest tertile as the reference. This analysis was adjusted for maternal age and parity.

In the later life study, association of parental with offspring obesity-adiposity was examined using multiple logistic regression. Reference category was non-overweight-obese parents, compared against one or both overweight-obese parents. Parental glycemic status included: maternal diabetes during pregnancy (yes/no) and paternal glucose intolerance at follow-up (yes/no). Birth weight and SES were also included in this analysis.

Given the long duration of the studies, some data were missing; we report number of available observations for each variable. We used SPSS version 21.0 (IBM corporation, Armonk, NY) for analysis.

### Ethics

All individual studies received approvals from KEMHRC Ethics Committee (DIP 2128/08-12-2021), IAEA-B12 (064/06-03-2006/15382/R0) and InDiaGDM (1333/1404/10-03-2014, (BT/IN/Denmark/02/CSY/2014)). InDiaGDM was also registered with ClinicalTrials.gov (NCT03388723). All adult participants signed an informed consent. For children below 18 years, we obtained parental consent, and children between 12 and 18 years also signed an informed assent.

## Results

In the neonatal study we included 816 diabetic pregnancies (74 type 1 diabetes, 102 type 2 diabetes, and 640 GDM) on whom all relevant data was available (maternal type of diabetes, gestational age at delivery, neonatal sex and weight), out of a total of 1057 pregnancies treated during this period. (Supplemental Fig. 1). Of the 241 excluded women data on age at conception, post-delivery BMI, and skinfolds, although available on less numbers, did not show a significant difference from that of included women (all p>0.05). Similarly, in the neonates there was no significant difference for sex ratio, gestational age, birth weight, abdominal circumference, and skinfolds (all p>0.05). (Supplemental Table S2). We used data on 372 NGT pregnancies for comparison.

In the later life study, we sent letters and called available phone numbers, and were able to contact 346 women treated for diabetes in pregnancy in our department. Of these 176 agreed to participate (133 were born in a GDM pregnancy, and 43 in a pre-gestational diabetic pregnancy (22 type 1 diabetes and 21 type 2 diabetes). Included women had similar age, BMI, glucose concentrations at diagnosis of GDM, and babies’ birth weight compared to the excluded (all p>0.05, Supplemental Table S3). Apart from the 176 offspring born in the index pregnancy, we also studied 24 siblings born in a subsequent diabetic pregnancy (20 were born in GDM, 3 in type 1 and 1 in type 2 diabetic pregnancies). Of these total 200 ODMs, 119 were < 10y and 81 were >=10y old. We also studied their parents (176 mothers and 150 fathers). In addition, we studied 177 ONDMs (93 < 10y and 84 >=10y old) and their parents (177 mothers, 163 fathers) as controls (Supplemental Fig.1).

### Neonatal Study

#### Mothers

Mothers with diabetes were older at conception, had higher parity and family history of diabetes compared to NGT mothers (Table 1). Within the diabetic group, mothers with type 1 diabetes were the youngest, thinnest and the least adipose; they also had longer duration of diabetes compared to the mothers with type 2 diabetes (median 8 y and 2y, respectively) and had the highest HbA1c (median, type 1 diabetes 8.5%, type 2 diabetes 7.0% and GDM 5.7%; p <0.05 for all). All mothers with type 1 diabetes received multiple daily injections of insulin; 50.8% with type 2 diabetes received insulin alone, 42.6% received insulin and OADs, and 3.3% received OADs alone. Of the GDM mothers, 62.0% received insulin alone, 20.8% received insulin along with OADs, 0.9% received OADs alone, and remaining were treated with lifestyle adjustment. Preterm and caesarean deliveries were more common in diabetic compared to non-diabetic pregnancies, highest in the mothers with type 1 diabetes.

**Table 1:** Maternal and neonatal characteristics in the neonatal study (DIP) according to maternal type of diabetes.

| Measurements | NGT |  | Type 1 diabetes |  | Type 2 diabetes |  | GDM |  | All (type 1+type 2+GDM) |  | P1 | P2 | P3 | P4 |
| --- | --- | --- | --- | --- | --- | --- | --- | --- | --- | --- | --- | --- | --- | --- |
|  | n | Median (25<sup>th</sup>, 75<sup>th</sup> p.) | n | Median (25<sup>th</sup>, 75<sup>th</sup> p.) | n | Median (25<sup>th</sup>, 75<sup>th</sup> p.) | n | Median (25<sup>th</sup>, 75<sup>th</sup> p.) | n | Median (25<sup>th</sup>, 75<sup>th</sup> p.) |  |  |  |  |
| **Mothers** |  |  |  |  |  |  |  |  |  |  |  |  |  |  |
| Age at conception (y) | 355 | 24.0 (20.8, 26.0) | 56 | 26.4 (23.0, 28.6) | 84 | 30.8 (27.5, 34.7) | 512 | 28.7 (25.8, 31.8) | 652 | 28.6 (25.7, 32.0) | **0.000** | **0.000** | **0.000** | **0.000** |
| Nulliparity |  | 229/364 (62.9 %) |  | 24/64 (37.5 %) |  | 24/91 (26.4 %) |  | 201/564 (35.6 %) |  | 249/719 (34.6 %) | **0.000** | **0.000** | **0.000** | **0.000** |
| Maternal height (cm) | 366 | 153.8 (150.3, 157.8) | 53 | 154 (150.2, 156.6) | 83 | 154.4 (150.6, 158.5) | 501 | 155 (150.9, 158.6) | 637 | 154.9 (150.6, 158.6) | 0.942 | 0.333 | **0.020** | **0.032** |
| Post-delivery weight (kg) | 331 | 54.1 (47.7, 60.7) | 50 | 61.2 (51.2, 68.6) | 78 | 65.1 (56.2, 77.5) | 487 | 64.1 (57.8, 72.7) | 615 | 64.0 (57.2, 72.6) | **0.000** | **0.000** | **0.000** | **0.000** |
| Post-delivery BMI (kg/m<sup>2</sup>) | 328 | 22.5 (20.1, 25.1) | 49 | 24.7 (22.3, 28.7) | 75 | 27.9 (23.8, 32.4) | 424 | 27.2 (24.9, 30.1) | 548 | 27.1 (24.5, 30.1) | **0.000** | **0.000** | **0.000** | **0.000** |
| Waist cir. (cm) | 164 | 76.5 (68.0, 83.5) | 42 | 89.9 (83.7, 95.7) | 64 | 96.3 (90.5, 102.4) | 352 | 96 (89.0, 102.0) | 458 | 95.4 (88.6, 102.0) | **0.000** | **0.000** | **0.000** | **0.000** |
| Sum of skinfolds (mm) | 164 | 49.8 (40.6, 69.4) | 43 | 86.3 (68.4, 107.5) | 64 | 101.7 (76.4, 130.1) | 374 | 101.2 (80.7, 122.1) | 481 | 99.2 (79.6, 121.9) | **0.000** | **0.000** | **0.000** | **0.000** |
| HbA1c (%) |  | - | 22 | 8.5 (6.7, 10.0) | 31 | 7.0 (6.1, 7.9) | 55 | 5.7 (5.3, 6.4) | 108 | 6.3 (5.6, 7.7) | - | - | - | - |
| HbA1c (mmol/mol) |  | - | 22 | 69.0 (50.0, 86.0) | 31 | 53.0 (43.0, 63.0) | 55 | 39.0 (34.0, 46.0) | 108 | 45.0 (38.0, 61.0) | - | - | - | - |
| **Neonates** |  |  |  |  |  |  |  |  |  |  |  |  |  |  |
| Male offspring |  | 185 (49.7) |  | 46 (62.2) |  | 52 (51.0) |  | 360 (56.3) |  | 458 (56.1) | 0.051 | 0.823 | **0.045** | **0.040** |
| Gestational age at delivery (weeks) | 372 | 39.1 (38.0, 40.0) | 74 | 37.3 (35.4, 38.1) | 102 | 37.5 (36.2, 38.3) | 640 | 38 (36.7, 39.1) | 816 | 37.8 (36.5, 39.0) | **0.000** | **0.000** | **0.000** | **0.000** |
| Birth Weight (kg) | 372 | 2.73 (2.50, 3.00) | 74 | 2.81 (2.30, 3.24) | 102 | 2.99 (2.47, 3.31) | 640 | 2.89 (2.52, 3.22) | 816 | 2.89 (2.50, 3.24) | **0.034** | **0.000** | **0.000** | **0.001** |
| Birth length (cm) | 343 | 48.2 (47.0, 49.5) | 48 | 49.0 (46.0, 50.3) | 72 | 49.1 (47.0, 50.0) | 413 | 48.9 (47.1, 50.2) | 533 | 49.0 (47.0, 50.2) | 0.388 | **0.015** | **0.005** | 0.057 |
| Ponderal index (g/cm<sup>3</sup>) | 343 | 2.41<br>(2.2, 2.6) | 48 | 2.57<br>(2.4, 2.7) | 72 | 2.56<br>(2.3, 2.7) | 413 | 2.49<br>(2.3, 2.7) | 533 | 2.5<br>(2.3, 2.7) | 0.064 | 0.140 | **0.008** | **0.028** |
| Abdominal circumference (cm) | 344 | 29.5<br>(28.1, 30.6) | 49 | 30.6<br>(28.8, 32.1) | 71 | 30.2<br>(28.1, 32.0) | 416 | 30<br>(28.5, 31.5) | 536 | 30.1<br>(28.5, 31.6) | **0.000** | **0.001** | **0.000** | **0.000** |
| Sum of skinfolds (mm) | 343 | 7.8<br>(6.8, 8.9) | 49 | 10.2<br>(8.7, 12.9) | 68 | 10.5<br>(8.2, 12.6) | 413 | 9.4<br>(8.0, 11.4) | 530 | 9.7<br>(8.0, 11.8) | **0.000** | **0.000** | **0.000** | **0.000** |

Data for Normal glucose tolerant (NGT) pregnancies was obtained from IAEA B12 and InDiaGDM Studies. Data on diabetic pregnancies was obtained from the clinical records of the Diabetes Unit KEM Hospital, Pune.
Values are median (25<sup>th</sup> and 75<sup>th</sup> percentiles) for continuous variables and n (%) for categorical variables.
p1: Type 1 Diabetes Vs Normal glucose tolerant, p2: Type 2 Diabetes Vs Normal glucose tolerant, p3: Gestational Diabetes Mellitus Vs Normal glucose tolerant, p4: All Vs Normal glucose tolerant

#### Neonates

As a group, ODM had higher birth weight, length, ponderal index, abdominal circumference and skin fold thickness compared to ONDM (Table 1). ODM had lower prevalence of SGA (21.8% vs. 42.9%) and higher prevalence of LGA (12.5% vs. 0.3%) as compared to ONDM. Offspring of mothers with type 1 diabetes had the highest ponderal index, abdominal circumference and skin fold thickness; offspring of mothers with type 2 diabetes and GDM followed in the order. (Fig. 1)

**Fig. 1.**
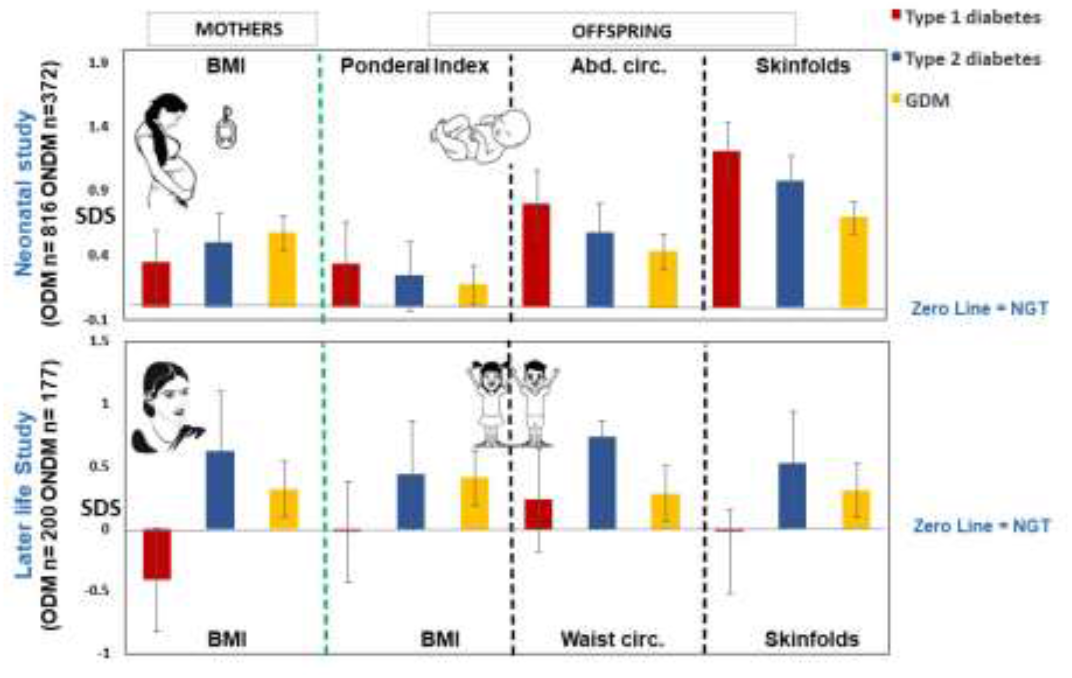
Obesity-adiposity in ODMs at birth and in later life. Figure shows obesity-adiposity indices in offspring of mothers with diabetes, at birth and in later life according to maternal type of diabetes. Number of observations for each category are shown in table 1 and 2. Data from normal glucose tolerant pregnancies is used as reference to calculated SD scores (age, sex, pubertal stage adjusted). BMI: Body Mass Index Abd. Cir. Abdominal circumference; Waist Circ.: waist circumference.

### Later life study

#### Mothers

As a group, mothers who were diabetic during pregnancy were heavier than mothers who were not diagnosed diabetic during pregnancy. Mothers with type 1 diabetes continued to be the thinnest (even in comparison to mothers who were not diabetic during pregnancy), mothers with type 2 diabetes were the most overweight-obese and centrally obese, GDM mothers were intermediate (Table 2, Fig. 1).

**Table 2:** Parental and offspring measurements in later life study (InDiaGDM) according to maternal type of diabetes in pregnancy.

|  | Maternal measurements at follow-up |  |  |  |  |  |  |  |  |
| --- | --- | --- | --- | --- | --- | --- | --- | --- | --- |
| Measurements | No diabetes<br>(n=177) | Type 1 diabetes<br>(n=22) | Type 2 diabetes<br>(n=21) | Gestational<br>Diabetes<br>(n=133) | All diabetes<br>(n=176) | p1 | p2 | p3 | p4 |
| Age (years) | 35.1 (31.1-39.6) | 33.6 (30.1-41.5) | 37.4 (33.8-39.7) | 37.8 (34.6-43.1) | 37.4 (34.2-42.8) | 0.557 | 0.348 | **0.000** | **0.001** |
| Height (cm) | 155.0 (151.5-158.2) | 156.1 (150.7-161.7) | 155.0 (152.1-160.0) | 154.4 (150.4-156.3) | 154.5 (150.5-157.5) | 0.260 | 0.644 | **0.031** | 0.210 |
| Weight (kg) | 60.7 (54.1-70.2) | 56.6 (52.5-65.2) | 67.4 (61.1-80.4) | 63.3 (56.8-71.6) | 62.8 (56.7-71.5) | 0.229 | **0.004** | **0.018** | **0.018** |
| BMI (Kg/m<sup>2</sup>) | 25.8 (22.7-28.7) | 23.6 (22.01-25.5) | 28.2 (25.7-31.9) | 27.1 (24.2-30.1) | 26.9 (23.9-29.7) | 0.070 | **0.003** | **0.001** | **0.002** |
| Waist (cm) | 89.1 (82.3-97.3) | 83.7 (80.9-91.0) | 96.8 (93.2-106.9) | 91.2 (58.7-98.7) | 91.2 (85.0-98.7) | 0.129 | **0.000** | **0.001** | **0.001** |
| Hip (cm) | 101.4 (95.0-107.9) | 98.3 (96.9-101.0) | 105.5 (100.2-115.9) | 103.3(95.7-111.7) | 102.4 (96.3-110.8) | 0.242 | **0.000** | **0.001** | **0.001** |
| Waist hip ratio | 0.88 (0.83-0.91) | 0.85 (0.83-0.89) | 0.92 (0.89-0.97) | 0.89 (0.85-0.93) | 0.89 (0.85-0.94) | 0.378 | **0.000** | **0.001** | **0.000** |
| Central obesity, n (%)<br>(Waist hip ratio >0.8) | 149 (86.6) | 18 (81.8) | 20 (95.2) | 122 (93.1) | 161 (92.0) | 0.776 | 0.175 | **0.047** | 0.105 |
| Overweight+ obesity, n (%) | 102 (57.6) | 8 (36.4) | 17 (81.0) | 91 (68.9) | 135 (67.8) | 0.065 | **0.034** | **0.041** | **0.000** |
|  | Offspring measurements |  |  |  |  |  |  |  |  |
| Measurements | No diabetes<br>(n=177) | Type 1 diabetes<br>(n=25) | Type 2 diabetes<br>(n=22) | Gestational<br>Diabetes<br>(n=153) | All diabetes<br>(n=200) | p1 | p2 | p3 | p4 |
| Age (years) | 9.4 (6.0-12.9) | 6.9 (4.4-15.3) | 5.1 (3.7-7.6) | 8.8 (5.1-12.9) | 8.1 (4.6-12.2) | 0.100 | **0.048** | 0.451 | 0.963 |
| SLI | 38.0 (34.0-42.8) | 37.5 (31.3-42.3) | 38.0 (34.0-41.0) | 40.0 (35.0-43.5) | 39.0 (35.0-43.0) | 0.449 | 0.399 | 0.223 | 0.464 |
| Birthweight (Kg) | 2.83 (2.50-3.25) | 2.70 (2.37-3.16) | 2.51 (2.46-3.09) | 2.85 (2.52-3.30) | 2.80 (2.50-3.25) | 0.124 | 0.050 | **0.018** | **0.047** |
| Birthweight SD score | -0.15 (-0.82-0.69) | 0.16 (-0.60-0.55) | -0.23 (-0.84-0.53) | 0.11 (-0.48-0.89) | 0.11 (-0.53-0.79) | 0.158 | 0.066 | **0.027** | 0.066 |
| Height (cm) | 132.8 (113.6-152.9) | 114.9 (104.7-152.7) | 109.5 (97.7-124.2) | 131.6 (106.8-154.6) | 126.8 (105.1-152.9) | 0.237 | 0.305 | 0.181 | 0.489 |
| Weight (Kg) | 28.4 (17.9-45.2) | 20.0 (16.2-43.8) | 15.8 (12.8-27.9) | 29.8 (17.1-48.8) | 26.5 (15.8-45.9) | 0.058 | 0.068 | **0.022** | 0.095 |
| BMI (Kg/m<sup>2</sup>) | 16.2 (14.1-19.1) | 15.5 (13.9-18.1) | 14.3 (13.4-18.2) | 16.8 (14.5-21.1) | 16.2 (14.4-20.8) | **0.001** | **0.000** | **0.000** | **0.000** |
| Overweight+ obesity, n (%) | 25 (14.2) | 3 (12) | 5 (22.7) | 39 (26) | 47 (23.9) | 0.765 | 0.293 | **0.007** | **0.018** |
| Waist (cm) | 59.8 (50.6-72.7) | 54.6 (51.0-67.4) | 51.0 (47.7-67.1) | 61.2 (51.0-76.3) | 58.8 (50.5-74.7) | **0.037** | 0.193 | 0.087 | 0.297 |
| Hip (cm) | 70.9 (57.7-87.0) | 60.9 (55.4-85.3) | 53.9 (50.7-71.4) | 72.0 (56.4-89.8) | 69.4 (55.3-86.1) | 0.130 | 0.151 | 0.058 | 0.194 |
| Waist hip ratio | 0.86 (0.82-0.90) | 0.88 (0.85-0.92) | 0.92 (0.90-0.98) | 0.89 (0.83-0.94) | 0.89 (0.84-0.94) | 0.249 | 0.344 | 0.773 | 0.650 |
| Sum of skinfolds (mm) | 40.5 (25.6-74.8) | 32.7 (25.6-58.2) | 28.7 (24.5-83.4) | 44.8 (27.6-88.1) | 43.4 (27.0-84.6) | 0.086 | 0.152 | 0.054 | 0.168 |
| **Paternal measurements at follow-up** |  |  |  |  |  |  |  |  |  |
| **Measurements** | **No diabetes**<br>**(n=162)** | **Type 1 diabetes**<br>**(n=19)** | **Type 2 diabetes**<br>**(n=14)** | **Gestational**<br>**Diabetes**<br>**(n=115)** | **All diabetes**<br>**(n=148)** | **p1** | **p2** | **p3** | **p4** |
| Age (years) | 39.7 (36.2-45.2) | 36.8 (33.6-44.7) | 39.9 (37.0-41.6) | 43.1 (38.5-46.6) | 41.8 (37.5-45.9) | **0.002** | **0.003** | **0.001** | **0.021** |
| Height (cm) | 167.7 (163.9-173.5) | 168.7 (163.8-172.4) | 166.8 (163.9-171.6) | 168.4 (164.8-174.0) | 168.2 (164.6-173.2) | 0.594 | 0.381 | 0.496 | 0.716 |
| Weight (kg) | 72.0 (64.5-80.7) | 75.0 (60.8-84.9) | 65.9 (60.3-82.6) | 73.5 (67.1-84.8) | 73.2 (65.9-84.7) | 0.120 | **0.055** | **0.043** | 0.111 |
| BMI (Kg/m<sup>2</sup>) | 25.3 (23.2-27.6) | 25.9 (24.6-27.9) | 23.4 (22.2-29.1) | 25.8 (23.6-28.7) | 25.8 (23.5-28.7) | 0.223 | 0.120 | 0.063 | 0.114 |
| Overweight +obesity, n (%) | 89 (54.6) | 11 (57.9) | 6 (42.9) | 71 (61.2) | 88 (59.1) | 0.785 | 0.398 | 0.272 | 0.427 |
| Waist (cm) | 95.6 (89.3-101.5) | 97.0 (89.9-103.4) | 90.1 (81.8-107.7) | 97.1 (91.6-104.4) | 96.9 (90.1-104.4) | 0.223 | 0.097 | 0.062 | 0.158 |
| Hip (cm) | 98.3 (95.2-104.0) | 99.4 (93.5-103.0) | 94.3 (88.0-105.6) | 100.9 (95.3-106.4) | 100.2 (94.6-105.5) | 0.197 | **0.051** | 0.104 | 0.316 |
| Waist hip ratio | 0.96 (0.93-0.99) | 0.97 (0.92-1.01) | 0.95 (0.91-1.03) | 0.97 (0.93-1.00) | 0.97 (0.93-1.00) | 0.114 | 0.719 | 0.645 | 0.797 |
| Central obesity, n (%)<br>(Waist hip ratio >0.9) | 148 (91.4) | 16 (84.2) | 11 (78.6) | 103 (88.8) | 130 (87.2) | 0.312 | 0.120 | 0.476 | 0.240 |
| Glucose intolerance<br>(prediabetes + diabetes),<br>n (%) | 91 (55.8) | 12 (63.2) | 7 (50.0) | 80 (69.0) | 116 (67.4) | 0.542 | 0.674 | **0.026** | **0.055** |

Classification of pregnancies as non-diabetic and child's birthweight in this group were based on maternal recall, diabetic pregnancies data was available in clinic records.
Values are median (25<sup>th</sup>-75<sup>th</sup> centile) for continuous variables n (%) for categorical variables
Offspring birth weight SD score was calculated for the group by adjusting for gestational age at delivery and gender.
Parental overweight-obesity (BMI ≥ 25 Kg/m<sup>2</sup>), glucose intolerance in father (ADA criteria, pre-diabetes + diabetes)
p1: type 1 diabetes Vs no diabetes; p2: type 2 diabetes Vs no diabetes, p3: GDM Vs no diabetes, p4: All Vs no diabetes

Of 133 women diagnosed with GDM, eighty-one (61%) mothers were diabetic at follow-up (61 diagnosed before, 20 diagnosed at follow-up OGTT) and 32 (24%) were prediabetic. Of the control mothers, 12 (7%) were diabetic at follow-up (7 diagnosed before and 5 at follow-up) and 46 (27%) were diagnosed with prediabetes.

#### Fathers

Fathers of ODM and ONDM showed a similar prevalence of overweight-obesity (59% vs 54%, p>0.05) and central obesity (87% vs 91%, p>0.05). Fathers of ODM had a higher prevalence of prediabetes (49% vs 33%, p=0.004), but that of diabetes was similar in the two groups (17% vs 23%, p>0.05).

#### Offspring

Age of the participants varied from 2 to 26 years. As a group, ODM had higher weight, BMI, WHR, skinfolds and body fat percent compared to ONDM (Supplemental Fig. 2), more prominent in males (Supplemental Fig. 3). Offspring of mothers with type 1 diabetes had lowest BMI, WHR, sum of skinfolds and body fat percent (DXA); offspring of mothers with type 2 diabetes had the highest measurements, and offspring of GDM were intermediate (Table 2, Fig. 1).

### Associations of offspring obesity-adiposity

#### Neonatal study

Multiple logistic regression showed that the risk of obesity, abdominal obesity and adiposity was the highest in neonates born to mothers with type 1 diabetes, there was no independent association with maternal BMI. Abdominal obesity of the neonate was additionally associated with higher maternal parity (Table 3, Supplemental Fig. 4).

**Table 3:** Logistic regression analysis to investigate association of type of maternal diabetes and size with neonatal obesity-adiposity (DIP study), and of parental diabetes and overweight-obesity with offspring obesity-adiposity in later life (InDiaGDM study)

|  | **Generalized obesity**<br>(Ponderal index >85<sup>th</sup> c.) | **Adiposity**<br>(Skinfolds >85<sup>th</sup> c.) | **Abdominal obesity**<br>(Abd. circ. >85<sup>th</sup> c.) |
| --- | --- | --- | --- |
| **Neonatal study** |  |  |  |
| **Maternal exposures** |  |  |  |
| NGT (n= 303)<br>(Reference category) | 1 | 1 | 1 |
| Type 1 diabetes (n=37) | **4.5 (1.9- 10.8)** | **17.6 (6.6 - 45.7)** | **4.0 (1.7 - 9.5)** |
| Type 2 diabetes (n=58) | **3.6 (1.5 - 8.9)** | **11.7 (4.3 - 31.4)** | **2.6 (1.1 - 6.2)** |
| GDM (n=310) | **2.8 (1.5- 5.2)** | **6.7 (3.0 - 15.0)** | **2.7 (1.5 - 4.9)** |
| BMI tertile 1<br>(BMI < 23.5 kg/m<sup>2</sup>)<br>(Reference category) | 1 | 1 | 1 |
| BMI tertile 2<br>(BMI 23.5 to 27.6 kg/m<sup>2</sup>) | 1.1 (0.6- 2.1) | 1.0 (0.5 -1.9) | 1.3 (0.7- 2.3) |
| BMI tertile 3<br>(BMI> 27.6 kg/m<sup>2</sup>) | 1.2 (0.6 - 2.3) | 1.5 (0.8- 3.0) | 1.4 (0.7- 2.5) |
| Nulliparity<br>(Reference category) | 1 | 1 | 1 |
| Multiparity<br>(parity 1 or more) | 1.1 (0.7- 1.8) | 1.0 (0.6 - 1.7) | **1.7 (1.0 - 2.7)** |
| Age (yrs.) | 0.3 (0.1- 1.3) | 0.7 (0.2- 3.3) | 0.5 (0.1-2.0) |
| **Later life study** |  |  |  |
|  | **Overweight-obesity**<br>(IOTF + WHO) | **Adiposity**<br>(Sum of skinfold<br>SD score > 85<sup>th</sup> c.) | **Abdominal obesity**<br>(Waist circumference<br>SD score>85<sup>th</sup> c.) |
| **Overweight-obese Parents** |  |  |  |
| Neither (n=66)<br>One (n=125)<br>Both (n=115) | 1.00<br>**6.13 (1.37-27.34)**<br>**9.99 (2.26-44.18)** | 1.00<br>**10.88 (1.40-84.21)**<br>**15.75 (2.04-121.54)** | 1.00<br>**5.76 (1.29-25.71)**<br>**6.76 (1.50-30.40)** |
| **Maternal diabetes in pregnancy** |  |  |  |
| No (n=148)<br>Yes (n=158) | 1.00<br>1.54 (0.82-2.89) | 1.00<br>1.34 (0.68-2.65) | 1.00<br>1.13 (0.58-2.19) |
| **Paternal glucose** |  |  |  |
| **intolerance at follow-up** |  |  |  |
| No (n=120) | 1.00 | 1.00 | 1.00 |
| Yes (n=186) | 1.29 (0.66-2.53) | 1.14 (0.55-2.36) | 0.95 (0.47-1.90) |
| **Socio economic score** |  |  |  |
| Lower (n=206) | 1.00 | 1.00 | 1.00 |
| Higher (n=100) | 0.77 (0.39-1.49) | 0.62 (0.29-1.31) | 0.94 (0.47-1.87) |
| **Offspring birth weight SD score** |  |  |  |
| < Median (n=154) | 1.00 | 1.00 | 1.00 |
| > Median (n=152) | **1.96 (1.04-3.74)** | **2.03 (1.00-4.12)** | 1.45 (0.85-2.87) |

c.; centile.
Values are Odds ratio (95% Confidence intervals). Significant results are shown in bold.
The number of participants mentioned in the table represent those on whom all relevant data was available
Parental overweight-obesity ( $BMI \geq 25 \text{ Kg/m}^2$ ), glucose intolerance in father (ADA criteria, pre-diabetes + diabetes), maternal diabetes during pregnancy (yes or no)
Offspring birth weight SD score was calculated for the group by adjusting for gestational age at delivery and gender.
WHO-World Health Organization, IOTF: Internal obesity task force criteria

#### Later life study

On logistic regression, parental overweight-obesity had the most significant effect on offspring obesity-adiposity. When one parent was overweight-obese, their children had six times higher risk of being overweight-obese compared to those born to neither parent overweight-obese, the risk increased to ten times when both parents were overweight-obese (Table 3, Supplemental Fig. 4); the effect was higher in ODMs compared to ONDMs (Supplemental Fig. 5). Similar findings were observed for central obesity and adiposity. Child’s birth weight was also associated with overweight-obesity and adiposity but not with central obesity. In contrast to the neonatal study, maternal diabetes during pregnancy was not associated with child’s obesity-adiposity. Parental glycemic status and SES at follow up also were not associated.

### Heritability and Parent-of-origin effect

In later life study, mid-parental measures were significantly associated with corresponding offspring measurements (weight, height, BMI, waist, and hip circumferences) with beta coefficients (SD per SD) ranging between 0.2 and 0.4. These heritability estimates were similar in sons and daughters (Supplemental Table S5).

We next examined if there was a stronger association of maternal or paternal measurement with corresponding offspring trait (parent of origin effect). If maternal measurement shows a stronger association than paternal measurement, this would indicate a maternal effect and vice versa. We found significantly excess maternal influence for weight and waist circumference of the offspring. For sons, maternal effect was observed for waist circumference whereas for daughters, it was observed for weight and hip circumference (Supplemental Table S6). Thus, our data highlighted significant heritability, a parent of origin (maternal) effect as well as sex specificity.

## Discussion

Our results confirm that offspring born in Indian diabetic pregnancies are more obese-adipose at birth and in later life than those born in non-diabetic pregnancies. We found that determinants of offspring obesity-adiposity at birth and in later life are not the same. Our seminal observation is that the offspring of mothers with type 1 diabetes (the thinnest but the most hyperglycaemic during pregnancy) were the most obese-adipose at birth but the least obese-adipose in the later life, compared to the offspring born to the more obese-adipose but less hyperglycemic mothers with type 2 diabetes and GDM. Later life obesity-adiposity in the offspring was influenced by maternal and paternal overweight-obesity in an additive manner (Supplemental fig. 5), with some evidence of excess maternal effect (Supplemental Table S6). In the literature, it is tacitly assumed that *maternal diabetes (hyperglycemia) in pregnancy* is the causal factor for both neonatal and later life outcomes. This assumption is based predominantly on observational follow-up of ODM in western populations in whom obesity-adiposity is very prevalent in the pregnant mothers with diabetes (14), and therefore the individual contribution of these two related but distinct exposures is difficult to separate. Randomized controlled trials of intensive control of maternal diabetes in pregnancy show a reduction in offspring birth weight and LGA but no protection against later life obesity-adiposity, thus raising doubts about causal role of maternal *diabetes (hyperglycemia)* for *long-term* obesity-adiposity in the children (15, 16). Our observations in the relatively thinner Indian population provide clean evidence for difference in aetiology of these two outcomes and invite different preventive strategies.

Review of literature shows considerable heterogeneity in the reporting of exposures and outcomes in studies investigating effects of parental size and glycemia on offspring size at birth and in later life. Most studies are in the Western population and demonstrate that maternal pregnancy glycemia is associated with offspring obesity-adiposity at birth but not always in later life (26-30). The observations are reported both in the non-diabetic (26-28) and diabetic pregnancies (14, 29-33). In an earlier study on smaller number of GDM pregnancies, we found that ODMs had higher birth weight compared to ONDMs (2.93 vs. 2.81kg) and larger skinfold thickness (18). In a study from Mysore, India, ODMs (n=49) had higher birthweight (3.36 vs 2.96 kg) and skinfold thickness compared to ONDM, and female ODM continued to have higher adiposity in childhood (24, 25, 34).

Offspring born to mothers with both obesity and diabetes are usually the most obese-adipose. There is only an occasional attempt to study independent effects of maternal size and glycemia or their interaction (14, 31). Paternal effects are rarely studied but are important especially in the postnatal period (11, 12, 26). Another consideration is the association of birth weight (or LGA) with subsequent obesity, it does show a positive relationship (35, 36) but the interesting question ‘is this association driven by the component of birth weight influenced by parental size or glycemia’ has not been investigated.

Type of maternal diabetes is also a surrogate for degree of hyperglycemia. Mothers with type 1 diabetes are usually the most hyperglycemic and GDM mothers the least. Our finding that the offspring of mothers with type 1 diabetes are the most obese-adipose at birth are supported strongly in the literature, as is the finding that these offspring are the least obese-adipose in later life (36, 37). An exception was seen in the Kaiser Permanente study where children of mothers with type 1 diabetes were overweight-obese even in the later life (38, 39); these mothers were considerably more obese (28.5 kg/m^2^, 34.1% obese) compared to mothers in our study.

Taken together, these studies concur with our findings that maternal diabetes (hyperglycemia) is a driver of neonatal obesity-adiposity and that this may be exaggerated by parental obesity-adiposity. These observations are strongly supportive of the Pedersen’s ‘hyperglycemia-hyperinsulinemia’ (3) and Freinkel’s ‘fuel-mediated teratogenesis’ models (4). The fuels refer to glucose, lipids, and amino acids (40). Effect of maternal diabetes on offspring obesity-adiposity seems to wane off rapidly during childhood and it is replaced by parental obesity-adiposity, with some contribution by birth weight of the offspring. Based on these findings we construct a working model for life course evolution of offspring obesity-adiposity (Supplemental Fig. 6).

### Strengths and limitations

We report findings in substantial numbers of common subtypes of diabetes in pregnancy, treated in a single department with uniform protocols. The GDM and control mothers in the neonatal study were defined by a 75 g OGTT, and maternal HbA1c measurements were available in representative numbers. A wide range of obesity-adiposity measurements in the offspring were available allowing study of size and body composition. Mothers with diabetes showed a clear disassociation between glycemia and size i.e., the thinnest (type 1 diabetes) were the most hyperglycaemic. Paternal size and glycaemic data were available in later life study, offering an opportunity to define their role in a condition usually ascribed only to maternal diabetes.

The neonatal study consisted of data from a retrospective hospital-based cohort, and some information was not available in a proportion of pregnancies. Diagnostic criteria for GDM changed during the course of the study which may introduce some heterogeneity. However, this was true for both patients and controls. Data on gestational weight gain, serial HbA1c in different trimesters, infant breast feeding and paternal size and glycemia was not available. Later life study was not a longitudinal growth data but a cross sectional follow-up in relatively smaller number of participants. This was due to the difficulties in contacting them because of changes in address and phone numbers. However, maternal age, body size and glucose concentrations at diagnosis, and offspring birth weight were similar in those who were followed and not followed. The control group in the later life study was mostly classified based on maternal recall rather than glucose data but the low prevalence of glucose intolerance at follow-up in mothers argue against substantial misclassification.

Given the peculiar characteristics of mothers in our study, generalizability to other western studies may not be possible, but our findings provide a unique opportunity to investigate a vexing issue.

**In summary**, we confirm increased short and long-term risk of obesity-adiposity in Indian ODMs. Maternal hyperglycemia is the driver of offspring obesity-adiposity during intrauterine life while in later life it was driven by bi-parental overweight-obesity. Possible mechanisms for these associations include genetic factors, epigenetic programming by the maternal intrauterine environment, or influence of a shared familial environment. Future genetic and epigenetic studies will help better understand the basis of offspring obesity-adiposity in diabetic pregnancies. Our observations provide clues to preventive strategies to reduce neonatal and later life obesity-adiposity. Strict maternal glycaemic control in pregnancy will help reduce neonatal obesity-adiposity, while family-based lifestyle interventions in the offspring may help reduce childhood obesity-adiposity, notwithstanding genetic contributions during both time periods.

## Supporting information

ESM Index Tables & Figure DIP +ODM_27 April 2023

## Data Availability

Data can be requested from Prof. C.S.Yajnik by applying with a 200 word plan of analysis, data sharing is subject to KEMHRC Ethics Committee approval and permission of Health Ministry Screening Committee of Govt. of India.

## Acknowledgements

Later life follow-up in this study was part of the InDiaGDM study funded by the Department of Biotechnology, New Delhi, India (BT/IN/Denmark/02/CSY/2014).

The International Atomic Energy Agency, Vienna, Austria, provided financial support for the IAEA-B12 study (15382/R0)

We are grateful to Prof. Patrick Catalano and Prof. Edwin Gale for informative discussions. We also acknowledge Dr. Meenakumai, Dr. Shailaja Kale, Pallavi Yajnik, Rasika Ladkat, Himangi Lubree, Sonali Rege, Rucha Wagh, Neelam Memane, Aboli Bhalerao, Vidya Gokhale, Swati Alekar, Alma Baptist, Preeti Kalel, Ajay, Abhay, Dr. Rohan Shah, Rupali Joshi and Shivani Rangnekar for their contributions to the study.

## Author contributions

SSW, KK, GRC, DSB and CSY were involved in study conceptualization; SSW, SRO, SSA, SND, HSD in subject recruitment, conduct of the study and treatment; SSW, SDJ, MKD were involved in data curation and analysis; DSB, DAR, RPK and SGW for laboratory analysis; SSW, SDJ, SBP and CSY in visualization, writing, and reviewing the manuscript.

## Data sharing statement

Prof. C.S.Yajnik is the guarantor of this work and, as such, had full access to all the data in the study and takes responsibility for the integrity of the data and the accuracy of the data analysis. Data can be requested from Prof. C.S.Yajnik by applying with a 200 word plan of analysis, data sharing is subject to KEMHRC Ethics Committee approval and Govt. of India’s Health Ministry Screening Committee permission.

## Declaration of interest

CSY is a visiting professor at the Danish Diabetes Academy (supported by Novo Nordisk, Denmark) and University of Southern Denmark during the conduct of the study and writing of this article.

None of the authors declare any conflict of interest.

Aspects of this work have been presented at the following national and international conferences: RSSDI (2021), DOHaD (2022), ObesityWeek (2022), SNEHA (2023).

## References

1. International Diabetes Federation. IDF Diabetes Atlas, 10th edn. Brussels, Belgium: 2021. Available at: https://www.diabetesatlas.org.

2. Krishnaveni GV, Yajnik CS. Foetal programming in a diabetic pregnancy: long-term implications for the offspring. Current Science. 2017 Oct 10:1321–6.

3. Pedersen J. Weight and length at birth of infants of diabetic mothers. European Journal of Endocrinology. 1954;16(4):330–42.

4. Freinkel N. Of pregnancy and progeny. Diabetes. 1980; 2:1023–1035.

5. Dabelea D, Pettitt DJ. Intrauterine diabetic environment confers risks for type 2 diabetes mellitus and obesity in the offspring, in addition to genetic susceptibility. Journal of Pediatric Endocrinology and Metabolism. 2001;14(8):1085–92.

6. Pettitt DJ, Bennett PH, Knowler WC, Robert Baird H, Aleck KA. Gestational diabetes mellitus and impaired glucose tolerance during pregnancy: long-term effects on obesity and glucose tolerance in the offspring. Diabetes. 1985;34(Supplement_2):119–22.

7. Pettitt DJ, Nelson RG, Saad MF, Bennett PH, Knowler WC. Diabetes and obesity in the offspring of Pima Indian women with diabetes during pregnancy. Diabetes care. 1993;16(1):310–4.

8. Ehrenberg HM, Mercer BM, Catalano PM. The influence of obesity and diabetes on the prevalence of macrosomia. American journal of obstetrics and gynecology. 2004 ;191(3):964–8.

9. Catalano PM, Ehrenberg HM. The short-and long-term implications of maternal obesity on the mother and her offspring. BJOG: An International Journal of Obstetrics & Gynaecology. 2006;113(10):1126–33.

10. Pettitt DJ, Baird HR, Aleck KA, Bennett PH, Knowler WC. Excessive obesity in offspring of Pima Indian women with diabetes during pregnancy. New England Journal of Medicine. 1983;308(5):242–5.

11. Pettitt DJ, Knowler WC, Bennett PH, Aleck KA, Baird HR: Obesity in offspring of diabetic Pima Indian women despite normal birth weight. Diabetes Care. 1987; 10:76–80.

12. Dabelea D, Hanson RL, Lindsay RS, Pettitt DJ, Imperatore G, Gabir MM, Roumain J, Bennett PH, Knowler WC. Intrauterine exposure to diabetes conveys risks for type 2 diabetes and obesity: a study of discordant sibships. Diabetes. 2000;49(12):2208–11.

13. Schaefer-Graf UM, Pawliczak J, Passow D, Hartmann R, Rossi R, Buhrer CH, Harder T, Plagemann A, Vetter K, Kordonouri O. Birth weight and parental BMI predict overweight in children from mothers with gestational diabetes. Diabetes care. 2005;28(7):1745–50.

14. Josefson JL, Catalano PM, Lowe WL, Scholtens DM, Kuang A, Dyer AR, Lowe LP, Metzger BE. The joint associations of maternal BMI and glycemia with childhood adiposity. The Journal of Clinical Endocrinology & Metabolism. 2020;105(7):2177–88.

15. Gillman MW, Oakey H, Baghurst PA, Volkmer RE, Robinson JS, Crowther CA. Effect of treatment of gestational diabetes mellitus on obesity in the next generation. Diabetes care. 2010;33(5):964–8.

16. Landon MB, Rice MM, Varner MW, Casey BM, Reddy UM, Wapner RJ, Rouse DJ, Biggio Jr JR, Thorp JM, Chien EK, Saade G. Mild gestational diabetes mellitus and long-term child health. Diabetes care. 2015;38(3):445–52.

17. Seshiah V., Balaji V., Balaji MS., et al. Gestational diabetes mellitus in India.J Assoc Physicians India. 2004; 52 (9): 707–11.

18. Kale SD, Kulkarni SR, Lubree HG, Meenakumari K, Deshpande VU, Rege SS, Deshpande J, Coyaji KJ, Yajnik CS. Characteristics of gestational diabetic mothers and their babies in an Indian diabetes clinic. JAPI. 2005; 53:857–62.

19. Katre P, Bhat D, Lubree H, Otiv S, Joshi S, Joglekar C, Rush E, Yajnik C. Vitamin B12 and folic acid supplementation and plasma total homocysteine concentrations in pregnant Indian women with low B12 and high folate status. Asia Pacific journal of clinical nutrition. 2010;19(3):335–43.

20. Rao S, Yajnik CS, Kanade A, Fall CH, Margetts BM, Jackson AA, Shier R, Joshi S, Rege S, Lubree H, Desai B. Intake of micronutrient-rich foods in rural Indian mothers is associated with the size of their babies at birth: Pune Maternal Nutrition Study. The Journal of nutrition. 2001;131(4):1217–24.

21. Joglekar CV, Fall CH, Deshpande VU, Joshi N, Bhalerao A, Solat V, Deokar TM, Chougule SD, Leary SD, Osmond C, Yajnik CS. Newborn size, infant and childhood growth, and body composition and cardiovascular disease risk factors at the age of 6 years: the Pune Maternal Nutrition Study. International journal of obesity. 2007; 31(10):1534–44.

22. Cole TJ, Lobstein T. Extended international (IOTF) body mass index cut-offs for thinness, overweight and obesity. Pediatric obesity. 2012; 7(4):284–94.

23. Mercedes DO, Onyango WA, Borghi E, Siyam A, Nishida C, Siekmann J. Development of a WHO growth reference for school-aged children and adolescents. Bull World Health Organ. 2007;85(9):660–7.

24. Krishnaveni GV, Hill JC, Leary SD, Veena SR, Saperia J, Saroja A, Karat SC, Fall CH. Anthropometry, glucose tolerance, and insulin concentrations in Indian children: relationships to maternal glucose and insulin concentrations during pregnancy. Diabetes care. 2005; 28(12):2919–25.

25. Krishnaveni GV, Veena SR, Hill JC, Kehoe S, Karat SC, Fall CH. Intrauterine exposure to maternal diabetes is associated with higher adiposity and insulin resistance and clustering of cardiovascular risk markers in Indian children. Diabetes care. 2010 ;33(2):402–4.

26. Knight B, Shields BM, Hill A, Powell RJ, Wright D, Hattersley AT. The impact of maternal glycemia and obesity on early postnatal growth in a nondiabetic Caucasian population. Diabetes care. 2007; 30(4):777–83.

27. Tint MT, Sadananthan SA, Soh SE, Aris IM, Michael N, Tan KH, Shek LP, Yap F, Gluckman PD, Chong YS, Godfrey KM. Maternal glycemia during pregnancy and offspring abdominal adiposity measured by MRI in the neonatal period and preschool years: The Growing Up in Singapore Towards healthy Outcomes (GUSTO) prospective mother–offspring birth cohort study. The American journal of clinical nutrition. 2020;112(1):39–47.

28. West J, Santorelli G, Whincup PH, Smith L, Sattar NA, Cameron N, Farrar D, Collings P, Wright J, Lawlor DA. Association of maternal exposures with adiposity at age 4/5 years in white British and Pakistani children: findings from the Born in Bradford study. Diabetologia. 2018; 61(1):242–52.

29. HAPO Study Cooperative Research Group. Hyperglycemia and Adverse Pregnancy Outcome (HAPO) Study: associations with neonatal anthropometrics. Diabetes. 2009; 58(2):453–9.

30. Pettitt DJ, McKenna S, McLaughlin C, Patterson CC, Hadden DR, McCance DR. Maternal glucose at 28 weeks of gestation is not associated with obesity in 2-year-old offspring: the Belfast Hyperglycemia and Adverse Pregnancy Outcome (HAPO) family study. Diabetes care. 2010; 33(6):1219–23.

31. Lowe WL, Lowe LP, Kuang A, Catalano PM, Nodzenski M, Talbot O, Tam WH, Sacks DA, McCance D, Linder B, Lebenthal Y. Maternal glucose levels during pregnancy and childhood adiposity in the Hyperglycemia and Adverse Pregnancy Outcome Follow-up Study. Diabetologia. 2019; 62:598–610.

32. Bendor CD, Bardugo A, Rotem RS, Derazne E, Gerstein HC, Tzur D, Pinhas-Hamiel O, Tsur AM, Cukierman-Yaffe T, Lebenthal Y, Afek A. Glucose Intolerance in Pregnancy and Offspring Obesity in Late Adolescence. Diabetes Care. 2022; 45(7):1540–8

33. Catalano PM, Farrell K, Thomas A, Huston-Presley L, Mencin P, De Mouzon SH, Amini SB. Perinatal risk factors for childhood obesity and metabolic dysregulation. The American journal of clinical nutrition. 2009; 90(5):1303–13.

34. Hill JC, Krishnaveni GV, Annamma I, Leary SD, Fall CH. Glucose tolerance in pregnancy in South India: relationships to neonatal anthropometry. Acta obstetricia et gynecologica Scandinavica. 2005; 84(2):159–65.

35. Kaul P, Bowker SL, Savu A, Yeung RO, Donovan LE, Ryan EA. Association between maternal diabetes, being large for gestational age and breast-feeding on being overweight or obese in childhood. Diabetologia. 2019; 62(2):249–58.

36. Hammoud NM, Visser GH, van Rossem L, Biesma DH, Wit JM, de Valk HW. Long-term BMI and growth profiles in offspring of women with gestational diabetes. Diabetologia. 2018; 61(5):1037–45.

37. Pitchika A, Vehik K, Hummel S, Norris JM, Uusitalo UM, Yang J, Virtanen SM, Koletzko S, Andrén Aronsson C, Ziegler AG, Beyerlein A. Associations of maternal diabetes during pregnancy with overweight in offspring: results from the prospective TEDDY study. Obesity. 2018; 26(9):1457–66.

38. Wang X, Martinez MP, Chow T, Xiang AH. BMI growth trajectory from ages 2 to 6 years and its association with maternal obesity, diabetes during pregnancy, gestational weight gain, and breastfeeding. Pediatric obesity. 2020; 15(2): e12579.

39. Sidell M, Martinez MP, Chow T, Xiang AH. Types of diabetes during pregnancy and longitudinal BMI in offspring from birth to age 10 years. Pediatric Obesity. 2021; 16(8): e12776.

40. Kulkarni SR, Kumaran K, Rao SR, Chougule SD, Deokar TM, Bhalerao AJ, Solat VA, Bhat DS, Fall CH, Yajnik CS. Maternal lipids are as important as glucose for fetal growth: findings from the Pune Maternal Nutrition Study. Diabetes care. 2013; 36(9):2706–13.

