## Supplementary material for "Maternal diabetes influences neonatal obesity-adiposity but not in later life Offspring obesity in diabetic pregnancy": ESM Index Tables & Figure DIP +ODM_27 April 2023

**Supplemental material**

**Index**

| No. | Title | Page No. |
|---|---|---|
| **Supplemental tables** | | |
| S1 | Methodology of anthropometric measurements | 2-3 |
| S2 | Characteristics of the mothers and neonates who were included and excluded in the DIP study | 4 |
| S3 | Characteristics of the mothers who were followed and not followed up in later life study (InDiaGDM (Arm-3) study) | 5 |
| S4 | Offspring body composition (DXA) measurements according to type of maternal diabetes in later life study (InDiaGDM study) | 6 |
| S5 | Heritability estimates for anthropometric and glycemic traits. | 7 |
| S6 | Parent-of-origin effects of anthropometric traits | 8 |
| **Supplemental figures** | | |
| 1 | Participant flow chart | 9 |
| 2 | Comparison of anthropometric measurements of ODM with ONDM in later life study. | 10 |
| 3 | Sex wise differences for anthropometric and DXA measurements between ODM and ONDM. | 11 |
| 4 | Association of type of maternal diabetes with neonatal obesity, and of parental overweight-obesity with offspring overweight-obesity in later life | 12 |
| 5 | Overweight + obesity in offspring according to parental size in ODM and ONDM in later life study | 13 |
| 6 | Determinants of offspring obesity and adiposity at birth and in later life | 14 |

**Supplemental Table S1: Methodology of anthropometric measurements**

| | Site of measurement | Instrument and make | | Neonatal study | | Later life study | | |
|---|---|---|---|---|---|---|---|---|
| | | | Least count | Neonate | Mother | Offspring | Mother | Father |
| Birth weight | - | Electronic weighing scale (ATCO Healthcare Ltd, Mumbai, India) | 50 g | ✓ | X | ✓ | X | X |
| Crown heel length | - | Pedobaby (ETS J.M.B., Brussels, Belgium) | 0.1 cm | ✓ | X | X | X | X |
| Ponderal Index | Birth weight (gm)/ (length, $cm^3$) | - | - | ✓ | X | X | X | X |
| Weight | - | Electronic weighing scales (ATCO Healthcare Ltd, Mumbai, India) | 0.01 kg | X | ✓ | ✓ | ✓ | ✓ |
| Height | Head (Frankfurt plane) | Stadiometer (CMS Instruments Ltd, London, UK). | 0.1 cm | X | ✓ | ✓ | ✓ | ✓ |
| BMI | Weight (Kg)/ (height, $m^2$) | - | - | X | ✓ | ✓ | ✓ | ✓ |
| Abdominal circumference | Just above the umbilical cord | Non- stretchable fiberglass tape (CMS Instruments, London, UK) | 0.1 cm | ✓ | X | X | X | X |
| Waist circumference | Mid-point between lower border of the costal margin and the upper border of the iliac crest in the mid-axillary line | Non- stretchable fiberglass tape (CMS Instruments, London, UK) | 0.1 cm | X | ✓ | ✓ | ✓ | ✓ |
| Hip circumference | The greater trochanter (the widest portion of | Non- stretchable fiberglass tape (CMS Instruments, London, | 0.1 cm | X | ✓ | ✓ | ✓ | ✓ |

| | | | | | | | | |
|---|---|---|---|---|---|---|---|---|
| | the hip) | UK) | | | | | | |
| Skinfold thickness | | | | | | | | |
| Triceps | Mid-point between the tip of shoulder and tip of the olecranon process | Harpenden skinfold calipers (CMS Instruments, London, UK) | 0.1 mm | ✓ | ✓ | ✓ | X | X |
| Biceps | The anterior most bulging portion of the upper arm over the biceps | Harpenden skinfold calipers (CMS Instruments, London, UK) | 0.1 mm | X | ✓ | ✓ | X | X |
| Subscapular | Immediately below the inferior angle of the scapula | Harpenden skinfold calipers (CMS Instruments, London, UK) | 0.1 mm | ✓ | ✓ | ✓ | X | X |
| Suprailliac | Upper border of the iliac crest is palpated and marked with a pen in the mid-axillary line on the non-dominant side | Harpenden skinfold calipers (CMS Instruments, London, UK) | 0.1 mm | X | ✓ | ✓ | X | X |
| Total fat, lean mass and percentage body fat | | Dual X-ray absorptiometry (DXA) (Lunar DPX-IQ 240, Lunar Corporation, Madison, WI, USA) | - | X | X | ✓ | X | X |

**Supplemental Table S2: Characteristics of the mothers and neonates who were included and excluded in the DIP study**

| | Included (n=816) | Excluded (n=241) | p |
|---|---|---|---|
| **Mothers** | | | |
| Age at conception (years) | 28.6 (25.7, 32.0) | 29.6 (25.7, 32.8) | 0.285 |
| BMI ($Kg/m^2$) (post-delivery) | 27.1 (24.5, 30.1) | 27.3 (24.2, 32.2) | 0.625 |
| Sum of skinfolds (mm) (post-delivery) | 99.2 (79.6, 121.9) | 107.8 (89.7, 126.4) | 0.061 |
| **Neonates** | | | |
| Gestational age (weeks) | 37.8 (36.4, 39.0) | 36.5 (35.9, 38.7) | 0.067 |
| Birth weight g § | 2842.0 (± 636.1) | 2803.1 (± 636.5) | 0.530 |
| Abdominal circumference (cm) § | 30.1 (± 2.4) | 30.5 (± 2.9) | 0.275 |
| Sum of skinfolds (mm) | 9.7 (8.0, 11.8) | 9.9 (8.6, 11.2) | 0.892 |

§ Values are mean, (SD) for normally distributed variables or median ($25^{th}$ and $75^{th}$ percentiles) for skewed variables.

**Supplemental Table S3: Characteristics of the mothers who were followed and not followed up in later life study (InDiaGDM (Arm-3) study)**

| | **Followed (n=176)** | **Not followed (n=685)** | p |
|---|---|---|---|
| **Mothers** | | | |
| Age at conception (years) | 29.4 (4.7) | 29.5 (4.6) | 0.827 |
| Height (cm) | 154.3 (6.2) | 154.9 (6.3) | 0.433 |
| BMI (Kg/$m^2$) | 23.5 (21.3-26.6) | 24.6 (22.2-27.3) | 0.134 |
| FPG (mmol/L) | 5.4 (4.8-6.7) | 5.7 (4.8-6.8) | 0.484 |
| 2-hour glucose (mmol/L) | 9.1 (7.8-11.3) | 9.4 (8.2-11.2) | 0.379 |
| **Offspring** | | | |
| Birth weight (gm) | 2821.4 (641.3) | 2897.8 (639.5) | 0.191 |

Values are mean, (SD) for normally distributed variables or median ($25^{th}$ and $75^{th}$ percentiles) for skewed variables. BMI: Body mass index, FPG: Fasting plasma glucose.

**Supplemental Table S4: Offspring body composition (DXA) measurements according to type of maternal diabetes in later life study (InDiaGDM study)**

| **Measurements** | **No diabetes (n=168)** | **Type 1 diabetes (n=19)** | **Type 2 diabetes (n=23)** | **Gestational diabetes (n=138)** | **All diabetes (n=180)** | **p1** | **p2** | **p3** | **p4** |
|---|---|---|---|---|---|---|---|---|---|
| Age (years) | 9.4 (6.0-12.9) | 6.9 (4.4-15.3) | 5.1 (3.7-7.6) | 8.8 (5.1-12.9) | 8.1 (4.6-12.2) | 0.100 | **0.048** | 0.451 | 0.963 |
| Weight (Kg) | 28.4 (17.9-45.2) | 20.0 (16.2-43.8) | 15.8 (12.8-27.9) | 29.8 (17.1-48.8) | 26.5 (15.8-45.9) | 0.058 | 0.068 | **0.022** | 0.095 |
| Fat mass (Kg) | 6.5 (2.6-12.9) | 3.5 (2.6-10.8) | 2.3 (1.8-10.3) | 8.4 (2.7-16.6) | 7.0 (2.6-15.5) | **0.021** | **0.025** | **0.010** | **0.027** |
| Total fat % | 21.2 (13.9-32.5) | 18.9 (14.9-23.3) | 15.0 (11.5-36.5) | 26.2 (16.3-36.9) | 22.9 (15.1-34.6) | **0.019** | **0.016** | **0.005** | **0.012** |
| Lean mass (Kg) | 19.9 (14.4-29.2) | 15.5 (12.6-33.0) | 14.1 (1.2-19.8) | 20.3 (14.1-31.2) | 19.4 (13.4-29.7) | 0.332 | 0.484 | 0.243 | 0.499 |
| Total lean % | 75.0 (63.9-82.8) | 77.4 (73.1-81.9) | 81.6 (60.4-84.9) | 70.1 (60.0-80.3) | 73.5 (61.9-81.3) | **0.004** | **0.010** | **0.005** | **0.018** |

Median ($25^{th}$-$75^{th}$ centile) for continuous variables n (%) for categorical variables

p1: type 1 diabetes Vs no diabetes; p2: type 2 diabetes Vs no diabetes, p3: GDM Vs no diabetes, p4: All Vs no diabetes

Supplementary figure 2 shows age, gender and pubertal status standardized scores of these characteristics for statistical comparison.

**Supplemental Table S5**: Heritability estimates for anthropometric and glycemic traits.

| | **All Offsprings (N=309)** | |
|---|---|---|
| | **ß (SE)** | **p-value** |
| ***Anthropometric*** | | |
| *Height (cm)* | 0.242 (0.055) | <0.000 |
| *Weight (kg)* | 0.380 (0.052) | <0.000 |
| *BMI (kg/m$^2$)* | 0.400 (0.052) | <0.000 |
| *Waist (cm)* | 0.361 (0.053) | <0.000 |
| *Hip (cm)* | 0.374 (0.052) | <0.000 |
| | **Sons (N=186)** | |
| ***Anthropometric*** | **ß (SE)** | **p-value** |
| *Height (cm)* | 0.222 (0.069) | 0.002 |
| *Weight (kg)* | 0.328 (0.066) | <0.000 |
| *BMI (kg/m$^2$)* | 0.342 (0.066) | <0.000 |
| *Waist (cm)* | 0.312 (0.067) | <0.000 |
| *Hip (cm)* | 0.329 (0.066) | <0.000 |
| | **Daughters (N=123)** | |
| ***Anthropometric*** | **ß (SE)** | **p-value** |
| *Height (cm)* | 0.257 (0.093) | 0.004 |
| *Weight (kg)* | 0.460 (0.086) | <0.000 |
| *BMI (kg/m$^2$)* | 0.494 (0.086) | <0.000 |
| *Waist (cm)* | 0.425 (0.087) | <0.000 |
| *Hip (cm)* | 0.442 (0.087) | <0.000 |

Heritability is tested between mid-parental phenotype (predictor) and off-spring phenotype (outcome) using linear model and expressed as ß and corresponding p-values.

**Supplemental Table S6**: Parent-of-origin effects of anthropometric traits.

| ***All offsprings*** | **MC (N=348)** | | **FC (N=309)** | | |
|---|---|---|---|---|---|
| | **ß (SE)** | **p-value** | **ß (SE)** | **p-value** | **Z (p-value)** |
| *Height (cm)* | 0.161 (0.059) | 0.007 | 0.132 (0.059) | 0.021 | 0.35 (0.730) |
| *Weight (kg)* | 0.327 (0.053) | <0.000 | 0.154 (0.054) | 0.005 | **2.29 (0.022)** |
| *BMI (kg/m$^2$)* | 0.284 (0.052) | <0.000 | 0.233 (0.053) | <0.000 | 0.69 (0.492) |
| *Waist (cm)* | 0.314 (0.054) | <0.000 | 0.153 (0.053) | 0.005 | **2.13 (0.033)** |
| *Hip (cm)* | 0.282 (0.054) | <0.000 | 0.195 (0.054) | <0.000 | 1.14 (0.255) |
| ***Sons*** | **MC (N=207)** | | **FC (N=186)** | | |
| | **ß (SE)** | **p-value** | **ß (SE)** | **p-value** | **Z (p-value)** |
| *Height (cm)* | 0.165 (0.079) | 0.038 | 0.104 (0.075) | 0.187 | 0.56 (0.575) |
| *Weight (kg)* | 0.248 (0.067) | 0.001 | 0.163 (0.069) | 0.026 | 0.88 (0.377) |
| *BMI (kg/m$^2$)* | 0.214 (0.065) | 0.003 | 0.233 (0.067) | 0.001 | -0.20 (0.839) |
| *Waist (cm)* | 0.302 (0.067) | <0.000 | 0.113 (0.063) | 0.111 | **2.06 (0.040)** |
| *Hip (cm)* | 0.210 (0.070) | 0.004 | 0.219 (0.067) | 0.003 | -0.09 (0.926) |
| ***Daughters*** | **MC (N=141)** | | **FC (N=123)** | | |
| | **ß (SE)** | **p-value** | **ß (SE)** | **p-value** | **Z (p-value)** |
| *Height (cm)* | 0.147 (0.090) | 0.111 | 0.169 (0.090) | 0.067 | -0.17 (0.863) |
| *Weight (kg)* | 0.453 (0.085) | <0.000 | 0.139 (0.086) | 0.090 | **2.60 (0.009)** |
| *BMI (kg/m$^2$)* | 0.402 (0.088) | <0.000 | 0.222 (0.089) | 0.007 | 1.44 (0.150) |
| *Waist (cm)* | 0.308 (0.094) | 0.001 | 0.216 (0.101) | 0.016 | 0.67 (0.505) |
| *Hip (cm)* | 0.401 (0.091) | <0.000 | 0.143 (0.090) | 0.100 | **2.02 (0.044)** |

Association between offspring measures with that of each of the parents are presented as ß values and SE. Differences between maternal and paternal effects are presented as Z scores and p-values

**Supplemental Figure 1: Participant Flow chart**

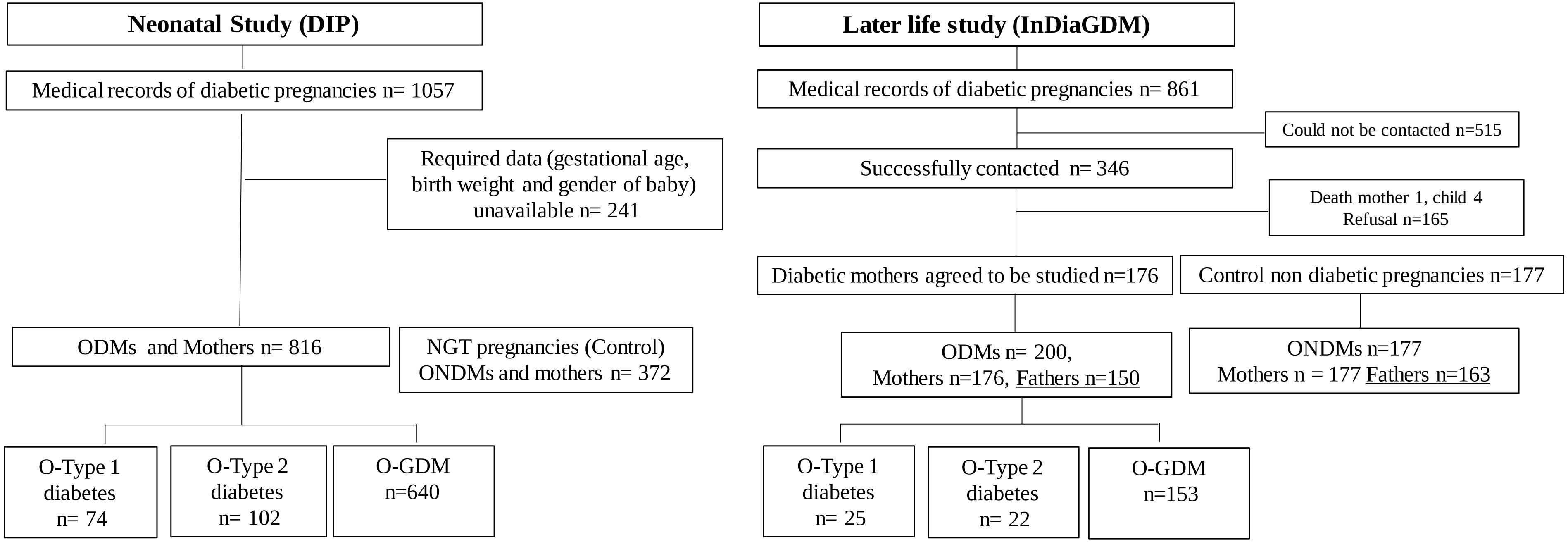

ODM: Offspring of mothers with diabetes in pregnancy
ONDM: Offspring of mothers without diabetes in pregnancy
NGT: Normal glucose tolerance
O-Type 1 diabetes: Offspring of mothers with type 1 diabetes
O-Type 2 diabetes: Offspring of mothers with type 2 diabetes

**Supplemental Figure 2: Comparison of anthropometric measurements of ODM with ONDM in later life study.**

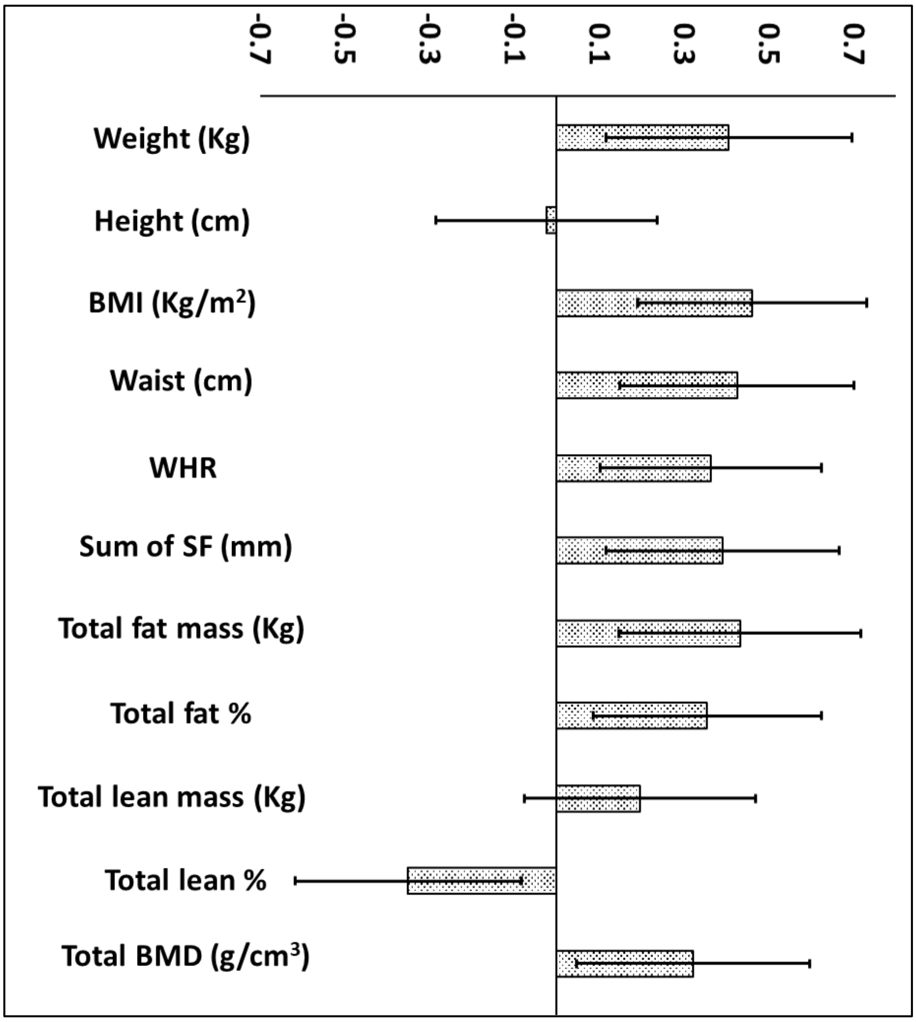

Mean SD differences (95% CI) for body size and composition (DXA) measurements of ODM (n=200) compared to ONDM (reference group, n=177) adjusted for age, sex, and pubertal stage. DXA measurements available on 181 ODM and 168 ONDM.
ODM= offspring of diabetic mothers, ONDM = offspring of non-diabetic mothers,
BMI = Body mass index, WHR = Waist hip ratio, SF: Skin fold, BMD= Bone mineral density

**Supplemental figure 3: Sex wise differences for anthropometric and DXA measurements between ODM and ONDM. Boys (n=121 ODM and 103 ONDM) and Girls (n=79 ODM and 74 ONDM) in later life study.**

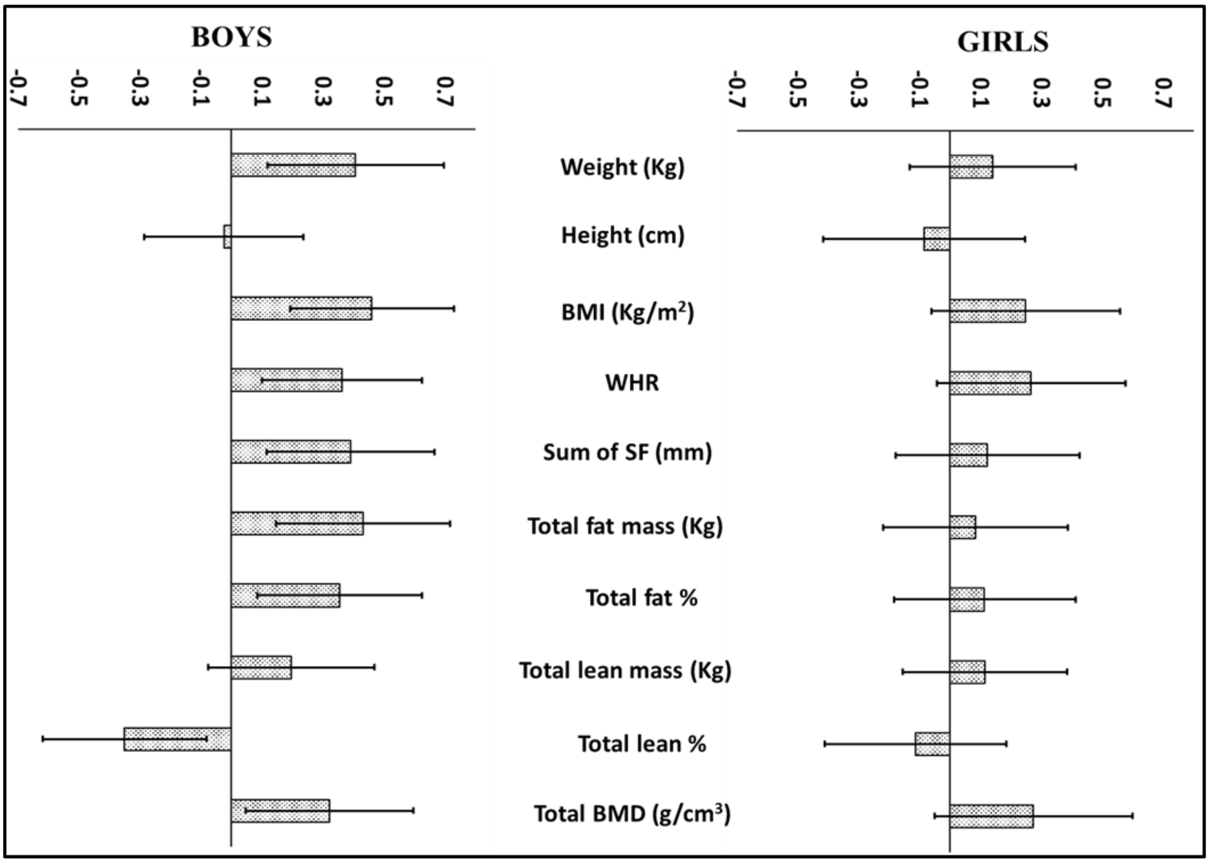

Mean SD differences (95% CI) for anthropometric and DXA measurements in ODM compared to ONDM, adjusted for age and pubertal stage. DXA measurements available in boys (100 ODM and 99 ONDM) girls (71 ODM and 69 ONDM)

**Supplemental figure 4: Association of type of maternal diabetes with neonatal obesity, and of parental overweight-obesity with offspring overweight-obesity in later life**

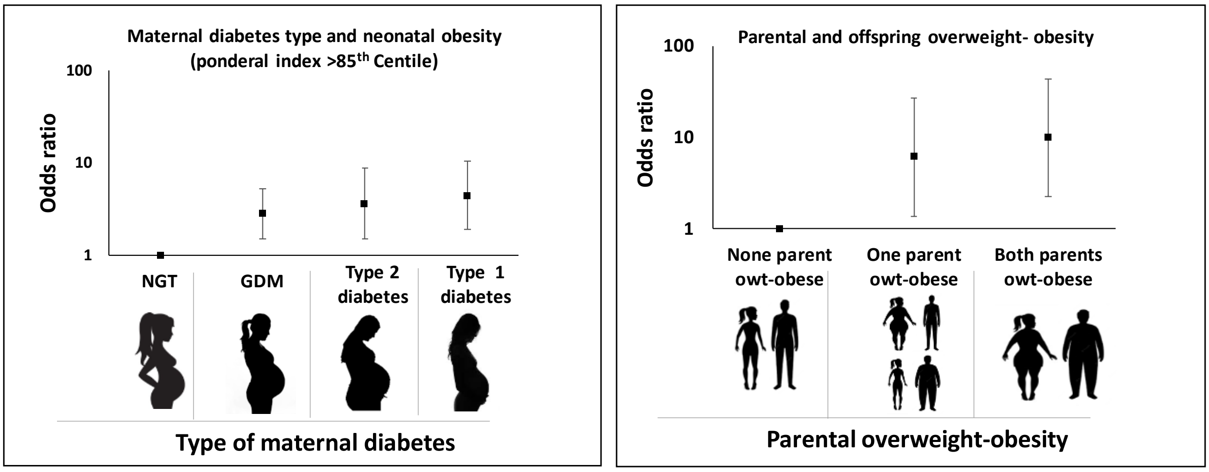

Figure based on data shown in table 3.

NGT: Normal glucose tolerant, GDM: Gestational diabetes mellitus. Parental overweight-obesity (WHO criteria, BMI >=25 Kg/m2). Offspring overweight-obesity in later life study: IOTF (2-18 years) and WHO criteria (>18 years).

**Supplemental figure 5: Overweight + obesity in offspring according to parental size in ODM and ONDM in later life study**

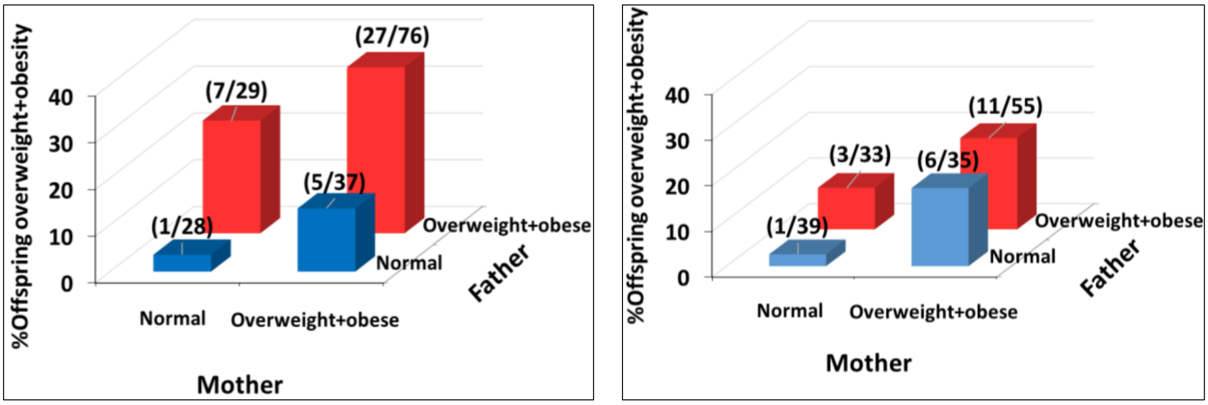

Figure shows association between parental and offspring overweight-obesity, data from diabetic and non-diabetic pregnancies is shown separately. There is a progressive increase in the offspring overweight-obesity when none, one or both parents are overweight-obese. The association is similar whether the offspring was born in a diabetic or a non-diabetic pregnancy, though proportion of overweight-obesity was higher in children born in diabetic pregnancies. The figure highlights biparental transmission of overweight-obesity in both diabetic and non-diabetic pregnancies.

Offspring: overweight-obesity were classified using IOTF (2-18 years) and WHO criteria (>18 years and parents). Overweight-obesity in parents (WHO criteria, BMI >=25 Kg/m2) was measured at follow up.

ODM: Offspring of diabetic mothers, ONDM: Offspring of non-diabetic mothers, IOTF: International Obesity Task Force, WHO: World Health Organization

**Supplemental figure 6: Determinants of offspring obesity and adiposity at birth and in later life**

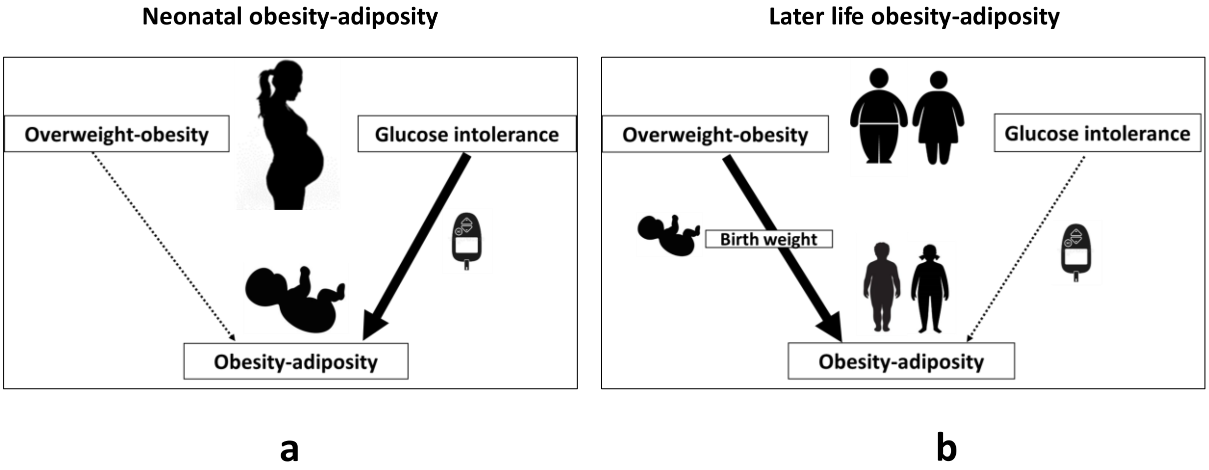

(a) Maternal diabetes promotes neonatal obesity-adiposity but not in later life.
(b) Bi-parental overweight-obesity promotes later life obesity-adiposity in the offspring
